# SARIMA Forecasts of Dengue Incidence in Brazil, Mexico, Singapore, Sri Lanka, and Thailand: Model Performance and the Significance of Reporting Delays

**DOI:** 10.1101/2020.06.26.20141093

**Authors:** Pete Riley, Michal Ben-Nun, James Turtle, David Bacon, Steven Riley

## Abstract

Timely and accurate knowledge of Dengue incidence is of value to public health professionals because it helps to enable the precise communication of risk, improved allocation of resources to potential interventions, and improved planning for the provision of clinical care of severe cases. Therefore, many national public health organizations make local Dengue incidence data publicly available for individuals and organizations to use to manage current risk. The availability of these data has also resulted in active research into the forecasting of Dengue incidence as a way to increase the public health value of incidence data. Here, we robustly assess time-series-based forecasting approaches against a null model (historical average incidence) for the forecasting of incidence up to four months ahead. We used publicly available data from multiple countries: Brazil, Mexico, Singapore, Sri Lanka, and Thailand; and found that our time series methods are more accurate than the null model across all populations, especially for 1-and 2-month ahead forecasts. We tested whether the inclusion of climatic data improved forecast accuracy and found only modest, if any improvements. We also tested whether national timeseries forecasts are more accurate if made from aggregate sub-national forecasts, and found mixed results. We used our forecasting results to illustrate the high value of increased reporting speed. This framework and test data are available as an R package. The non-mechanistic approaches described here motivates further research into the use of disease-dynamic models to increase the accuracy of medium-term Dengue forecasting across multiple populations.

**Author summary:** Dengue is a mosquito-borne disease caused by the Dengue virus. Since the Second World War it has evolved into a global problem, securing a foothold in more than 100 countries. Each year, hundreds of millions of people become infected, and upwards of 10,000 die from the disease. Thus, being able to accurately forecast the number of cases likely to emerge in particular locations is vital for public health professionals to be able to develop appropriate plans. In this study, we have refined a technique that allows us to forecast the number of cases of Dengue in a particular location, up to four months in advance. We test the approach using state-level and national-level data from Brazil, Mexico, Singapore, Sri Lanka, and Thailand. We found that the model can generally make useful forecasts, particularly on a two-month horizon. We tested whether information about climatic conditions improved the forecast, and found only modest improvements to the forecast. Our results highlight the need for both timely and accurate reports. We also anticipate that this approach may be more generally useful to the scientific community; thus, we are releasing a framework, which will allow interested parties to replicate our work, as well as apply it to other sources of Dengue data, as well as other infectious diseases in general.

## Introduction

Forecasting the near- and long-term evolution of Dengue incidence within a country has obvious value for policy makers. Dengue is a mosquito-borne disease caused by the Dengue virus, affecting most tropical regions of the world [1]. Each year, between 50 and 500 million people are infected with Dengue. Of these between 10,000 and 20,000 people die [2, 3]. In spite of the disease being endemic, seasons vary dramatically from one to the next, sometimes by more than an order of magnitude [4]. Knowledge of the estimated total number of cases, the timing of the peak, and near-term incidence can allow public health personnel to allocate limited resources appropriately, particularly when Dengue may be competing with other diseases. Accurate predictions of large increases in incidence would allow health care managers to prepare for a surge of patients, as well as more proactive interventions, such as vector control.

A number of statistical and mechanistic models have been developed with the aim of modeling or forecasting Dengue in various settings [5–13]. While ultimately, it is likely that mechanistic approaches [10, 14], should outperform statistical [15, 16] and machine learning (ML) approaches [17], our current understanding of the complex dynamics associated with vector-borne diseases, as well as the limitations in available data, suggest that statistical techniques should be considered first. Of the statistical approaches, Seasonal Autoregressive Integrated Moving Average (SARIMA) models have received the most attention [18–20]. Recently, these models have been applied in a pseudo-forecasting mode to assess their performance. In one study, the best overall performing model relied on lagged observations of one month, with three yearly lag terms and the first yearly difference [6], suggesting that there was a long term trend in the data and that the average of the last three years observations for a given month was a good model if adjusted by a single most recent observation from the current year. Notably, climate data did not appreciably improve the power of the SARIMA models.

In this study, we describe a statistical technique for predicting Dengue incidence rates from one to four months in the future using a family of SARIMA models. We apply the model to districts/provinces/states within five distinct countries (Brazil, Mexico, Singapore, Sri Lanka, and Thailand) for which reliable monthly or weekly data are available.

## Results

We first examined the data for evidence of seasonality. We then applied a reasonably exhaustive set of SARIMA models to these data, first without, and then with the addition of co-variate data. Next we explored the effects of direct versus aggregate forecasting, and finally, we investigated the effects of reporting time delays.

### Periodicity/Seasonality in the incidence data

There is a clear seasonal component to the incidence profiles for Brazil (data publicly available for the interval 2001-2012), Thailand (2007-2018), Mexico (1985-2017), and Singapore (2005-2019) (Fig. 1). For Sri Lanka (2010-2019), the picture is more complex. This is, at least in part, complicated by the fact that the peak values in 2017 were more than six times higher than the average peak values over the previous decade. To explore whether possible seasonal signatures exist, we applied a wavelet transform to district level data of Sri Lanka (Fig. 2). With the exception of Ratnapura, there is no evidence for a stable, annual peak (with a frequency of 12 months). On the other hand, there is some evidence for a sustained signal at 28-32 months that is present in all of the top-five districts (and most of the other 19 districts). In the right column we explore the idea that outbreaks spread out from the capital, Colombo (blue line in each panel), to other districts (red line in each panel) by plotting the average phase of the amplitude with a frequency of 10-14 months; however, we find no consistent evidence for a lead/lag.

**Fig 1.**
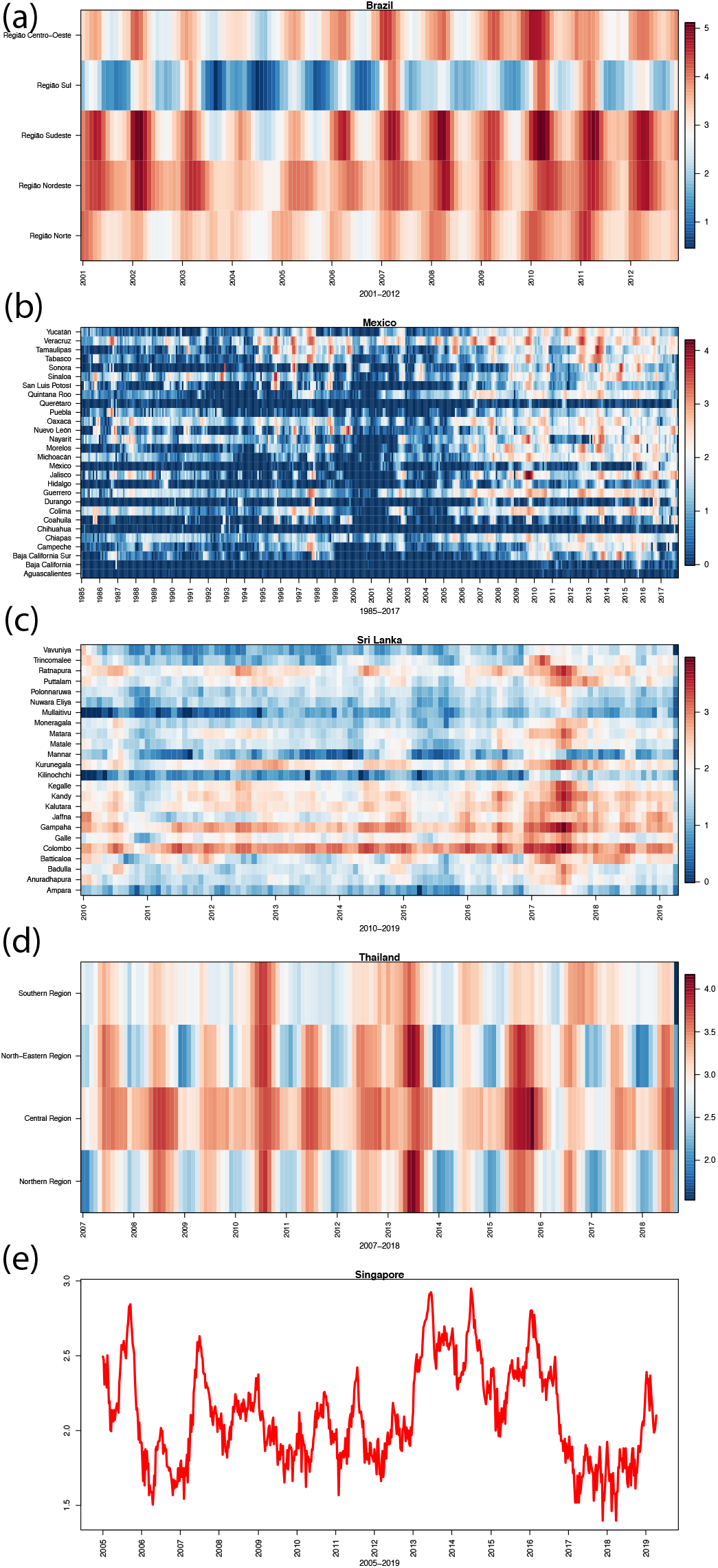
Heat maps for provinces or regions within (a) Brazil, (b) Mexico, (c) Sri Lanka and (d) Thailand. Monthly cadence is shown in all four panels. (e) Weekly national level incidence data for Singapore. In each panel, values represent *Log*^10^(*I* + 1), where *I* is the number of monthly or weekly cases in that region (or country).

**Fig 2.**
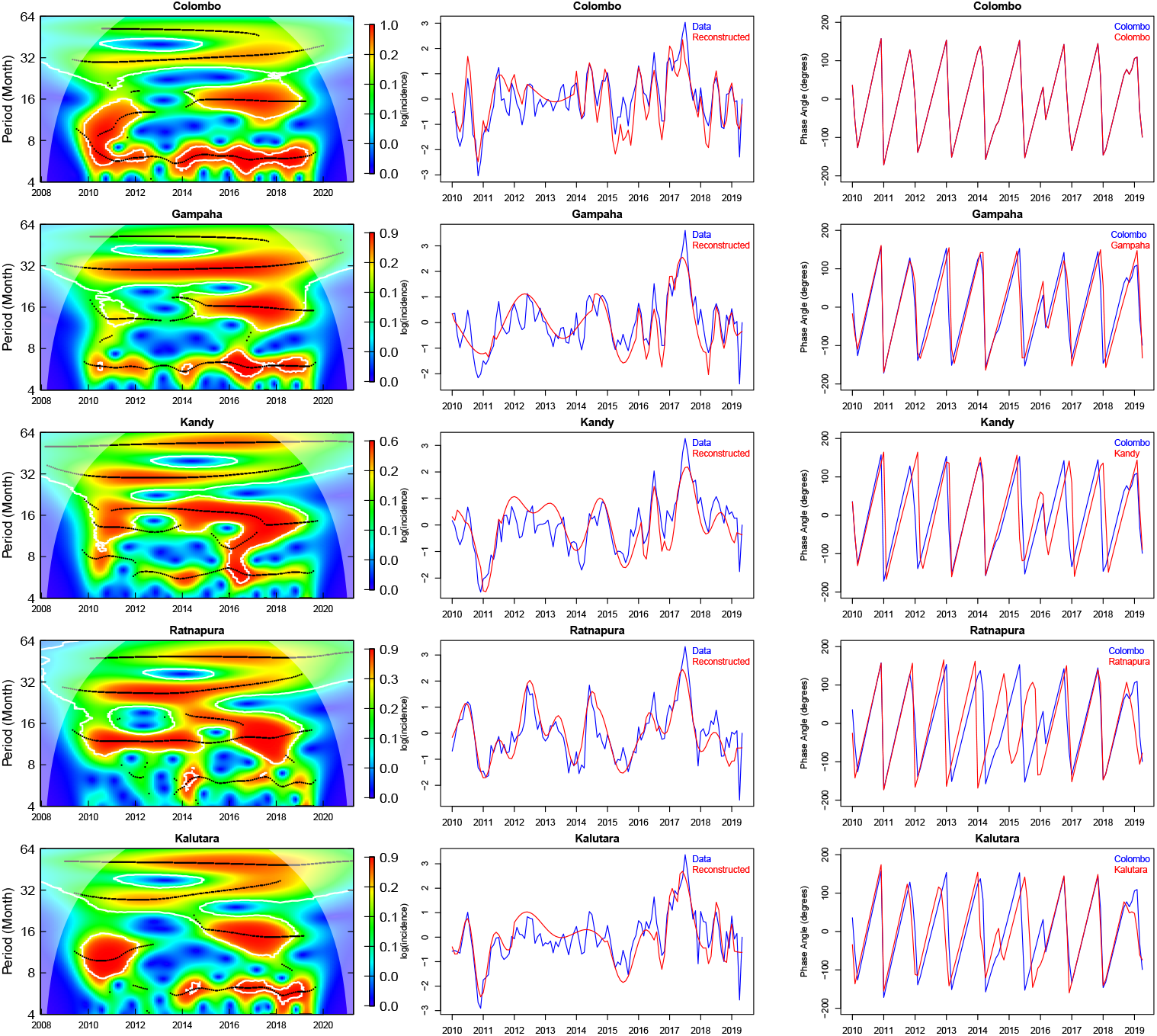
(Left) Wavelet analysis for Dengue in Sri Lanka at district level (top-five of 24 districts shown). (Middle) Original log incidence (blue) is compared with reconstructed data (red) from the wavelet transform. (Right) Average phase of the amplitudes with a frequency of 10-14 months. In each panel we compare a district (red) to Colombo (blue). The five districts are ordered by total number of cases with the capital Colombo having the largest number.

### SARIMA Analysis

Application of the leading eight SARIMA models to each of the states/provinces/regions within Brazil, Thailand, Mexico, and Singapore generally demonstrated that the (1, 0, 0)(3, 0, 0)^12^ SARIMA model performed best. For example, comparison of eight SARIMA models across 76 Thai provinces with our null historical model, showed that a simple monthly historical average with a 1-month-3-year lagged regression model of either the direct observations or their first difference (i.e., (1, 0, 0)(3, 0, 0)^12^ or (1, 0, 0)(3, 1, 0)^12^, respectively) performed best across all provinces (Fig. 3). Unsurprisingly, the Mean Absolute Error (MAE), tended to decrease moving from the most populous to least populous provinces, while the Mean Relative Absolute Error (MRAE) remained approximately constant from one province to another. Using the ratio of MAE(SARIMA) to MAE(NULL) as a measure of Skill Score (*SS*) for each SARIMA model, we infer that almost all eight of the SARIMA models outperformed the NULL (historical average) model. Finally, while the model skill decreased with increasing window of forecast (one month to four months), it is noteworthy that one-month forecasts are substantially better (*SS <* 0.5), and even four-month forecasts are still marginally better (*SS <* 1.0) than the historical average.

**Fig 3.**
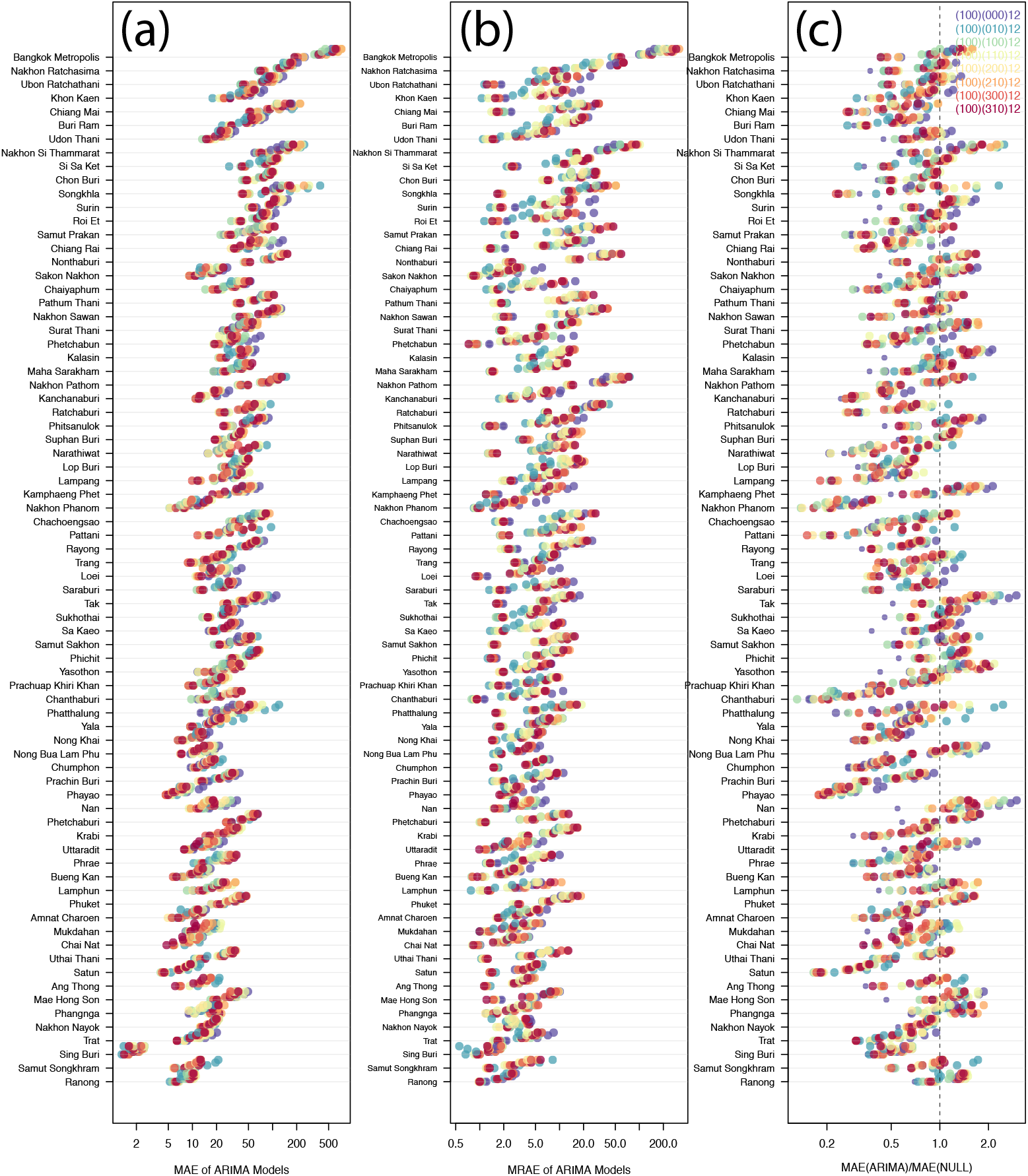
SARIMA analysis for the 76 Thailand provinces. (a) The mean absolute error (MAE) of the models; (b) the mean relative absolute error (MRAE) of the models; and (c) ratio of the MAE of the model to the MAE of the null model. The different colors denote the SARIMA models (see legend in top right panel), while dots of the same color, but shifted slightly upward in the y-direction show progressively longer-term predictions (one, two, three, and four months).

Brazilian Dengue incidence also displays a clear one-year modulation (Fig. 4). Forecasts using the 1-month-3-year model ((1, 0, 0)(3, 0, 0)^12^, SARIMA) generally perform well, with both the timing and amplitude of the peaks matching the observations. This remains true for two-month ahead forecasts (SI, Fig. S1), but degrades notably for three-month (SI, Fig. S2) and four-month (SI, Fig. S3) head forecasts. SARIMA forecasts for Mexico and Singapore are qualitatively similar to Brazil (Fig. 5). At both the state and country levels, the 1-month-3-year (1, 0, 0)(3, 0, 0)^12^ always performs best across all prediction horizons (one-four months). However, this result changes if we aggregate the state forecasts to predict the national values, rather than generating SARIMA forecasts based on the national data directly (see below).

**Fig 4.**
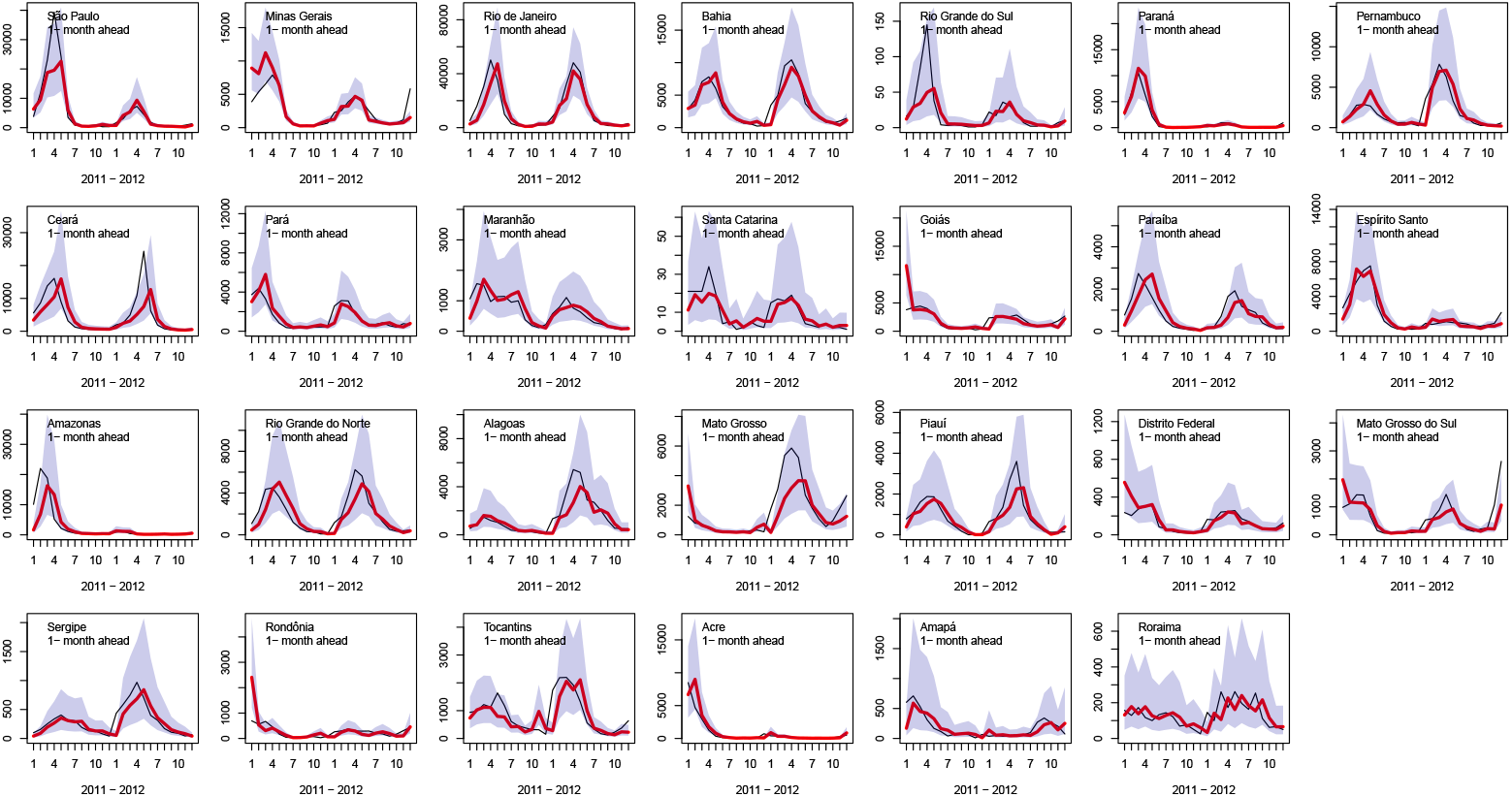
One-month ahead SARIMA predictions for Brazil Dengue incidence shown over a duration of two years. Each panel shows a different state (ordered by population from largest to smallest). The 1-month-3-year (1, 0, 0)(3, 0, 0)^12^ SARIMA model was used in each. Data are shown in black, predictions in red, and 5/95% CI are marked by the purple shaded regions.

**Fig 5.**
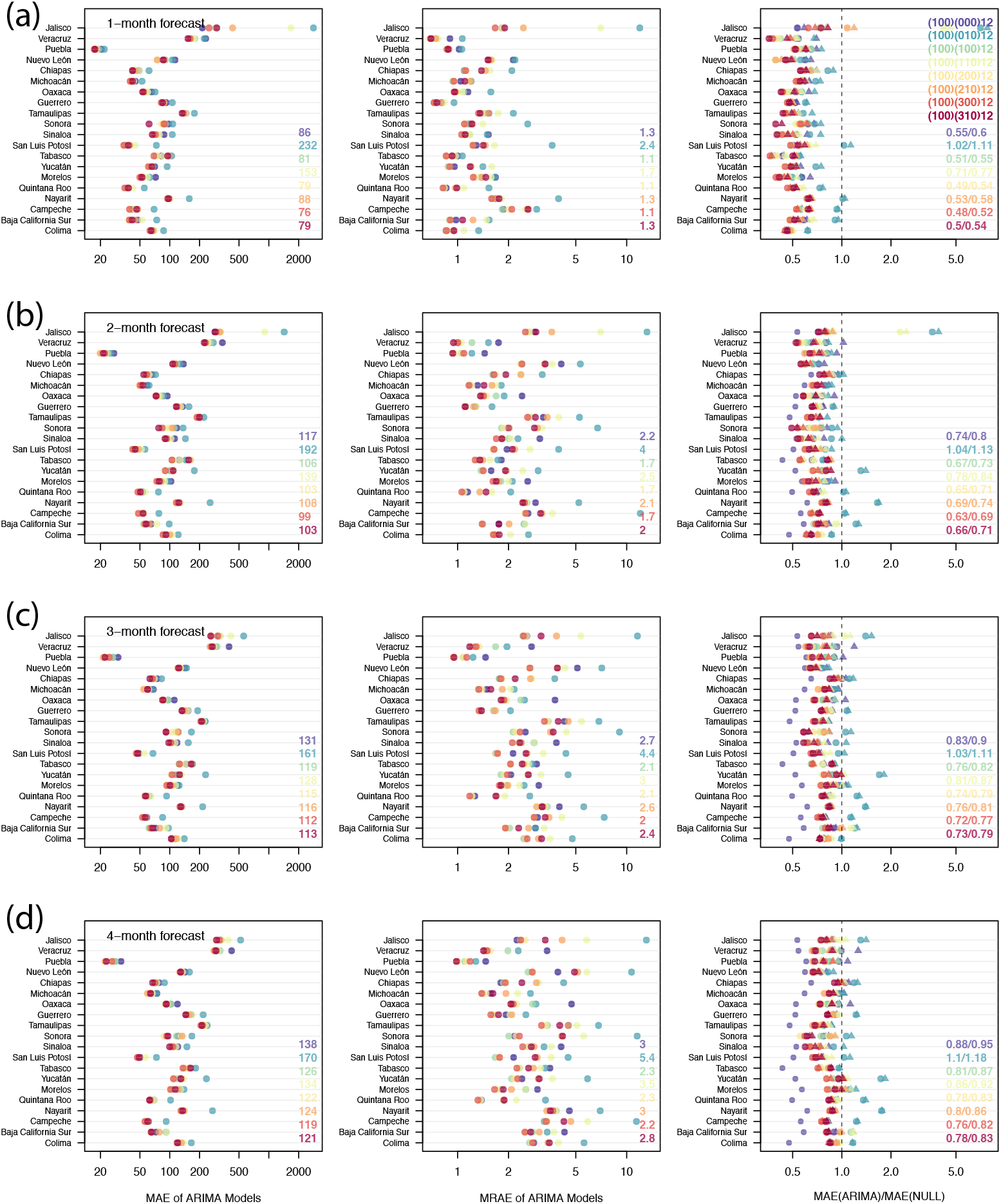
(a) One-, (b) two-, (c) three-, and (d) four-month ahead SARIMA predictions for Mexico Dengue incidence. The legend in each panel denotes the mean error of each SARIMA model. The list of SARIMA models are shown in the top legend of the top-right panel. See Fig 3 and text for more details.

### Inclusion of covariate parameters

Inclusion of covariate information, specifically precipitation, improved the forecast in some cases; however, even when this was the case, the effect was modest. Focusing first on Brazil, at both the state and regional level, the addition of the covariate information yielded a slightly more accurate forecast, which held for all windows from one to four months (Fig. 6). A similar analysis for Mexico also resulted in improvements at the state and national levels. For Thailand, while the covariate results were slightly better at the province level, they were essentially the same with or without the covariate at the national level. In all cases, where the covariate information improved the forecast, its effect was modest: it reduced the MAE by *∼* 10% at most. In a small number of cases, the results were more dramatic, with up to a 50% reduction in the MAE.

**Fig 6.**
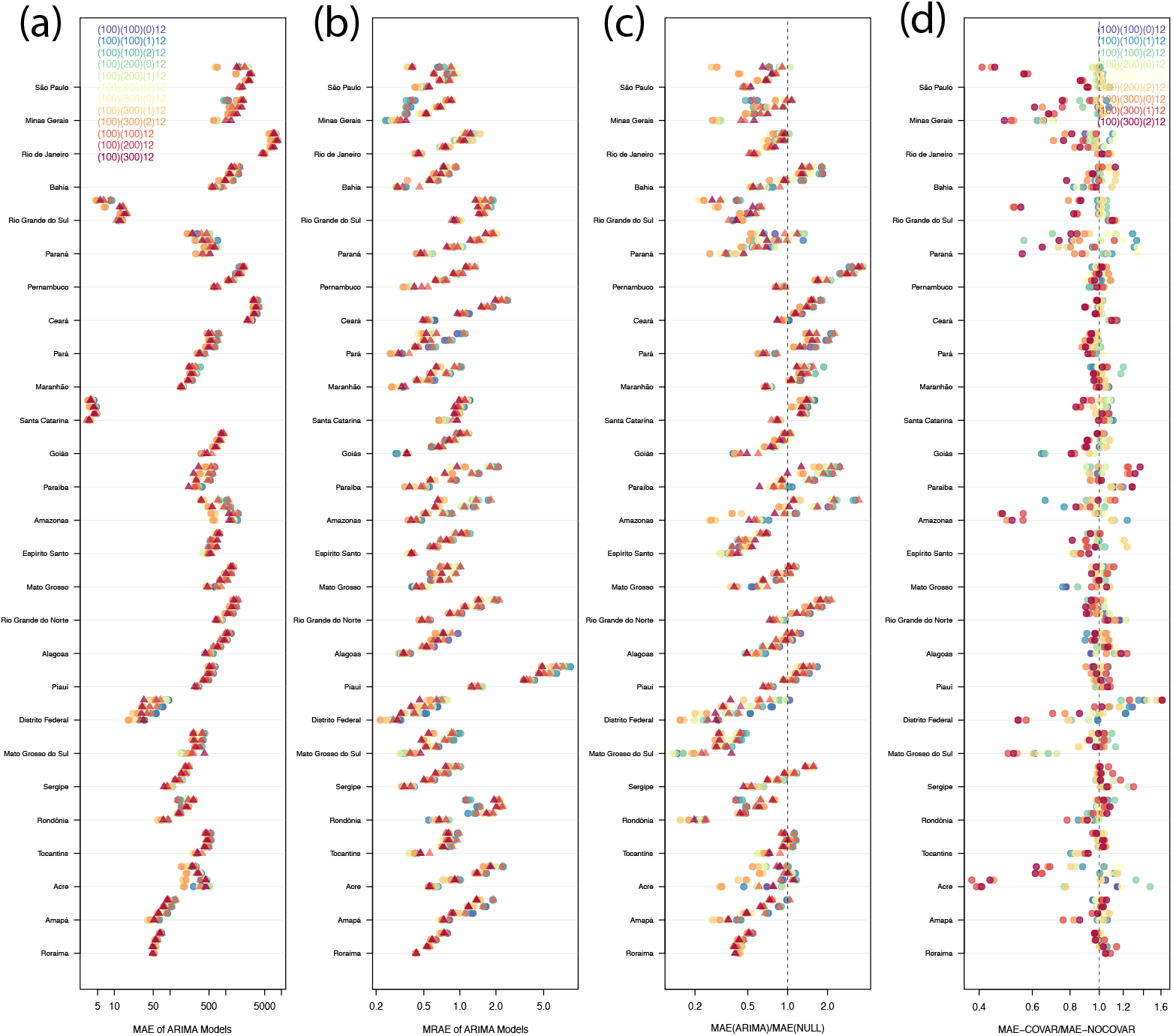
SARIMA Model results for Brazil, including the effects of a covariate. The format of the panels is the same as in Fig 3 for panels (a) - (c). (d) shows the MAE ratio when the precipitation covariate is/is not included. Triangles/circles indicate model results with/without the covariate.

Finally, we also explored the impact of covariate precipitation data for Singapore, for which there are weekly datasets (SI, Figs. S4 and S5). Again, we defined the null model to be the historical average of Dengue cases, this time calculated using weekly data, using the past ten years of data. We ran the same eight SARIMA models with forecast horizons ranging from 1 to 10 weeks. As the forecast horizon increased, the (1,0,0)(3,0,0) model performed best, with (1,0,0)(3,1,0) almost matching it. Here, however, seasonality appeared to play little role, presumably because the data were so irregular from one year to the next (Fig. 1(e)). The inclusion of precipitation data as a covariate (with lag times ranging from zero to four weeks) had no significant effect on the forecast.

### Direct versus Aggregated National Forecasting

Generally, we found that national forecasts were more accurate when made directly using the national data (the “direct” forecast), as opposed to forecasts assembled from individual region forecasts (the “aggregate” forecast). This effect, however, was location-dependent, and there were notable counterexamples.

For Mexico, the 1-month-3-year autoregressive models of either the actual observations or their first difference (i.e., (1, 0, 0)(3, 0, 0)^12^ and (1, 0, 0)(3, 1, 0)^12^) performed best for both direct and aggregate approaches (Fig. 7). Additionally, for forecast horizons of one and two months, the direct method produced better forecasts. For three and four month forecasts, at least for the most accurate SARIMA models, there was less separation between the direct and aggregate approaches.

**Fig 7.**
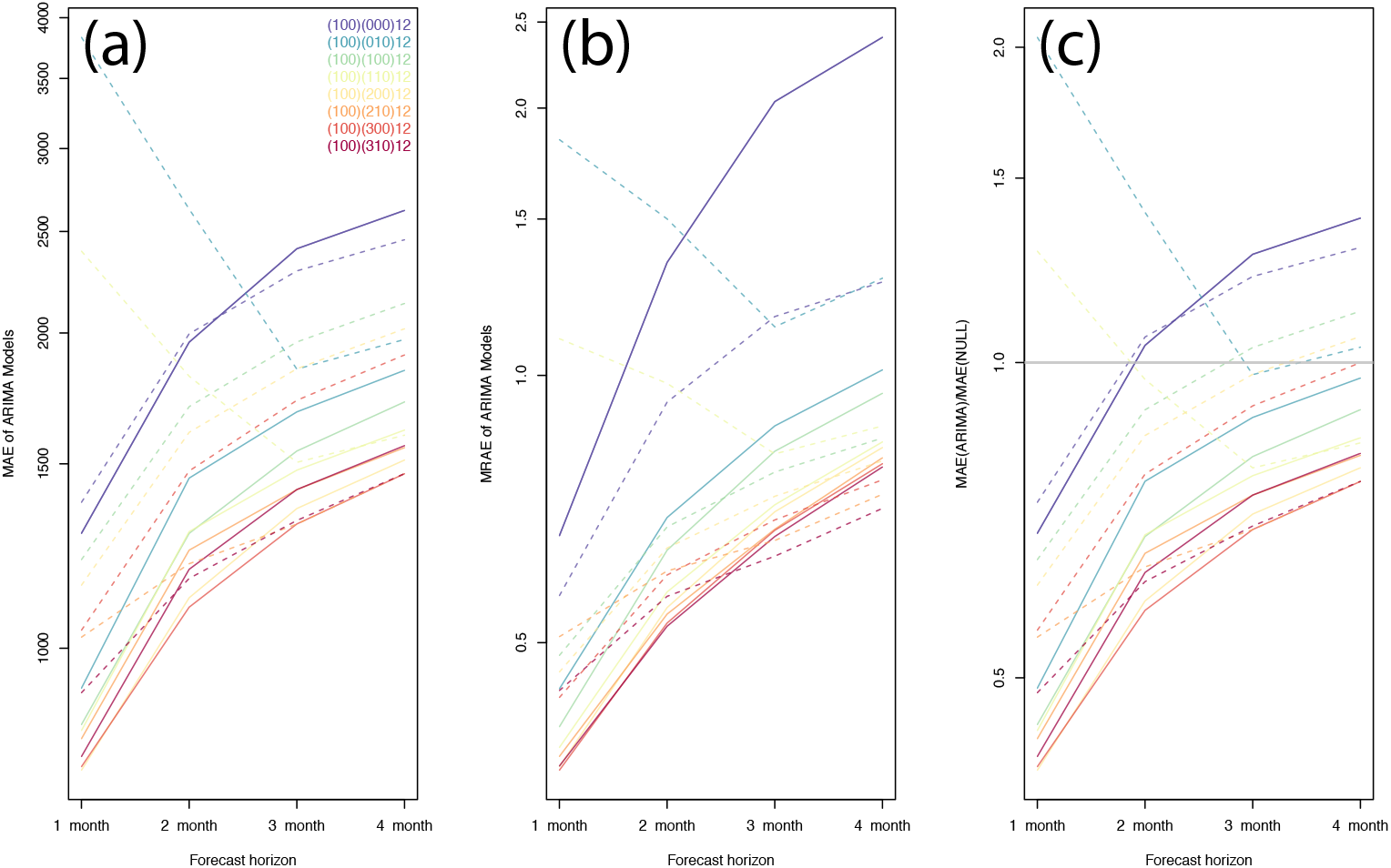
One, two, three, and four month ahead SARIMA predictions for Mexico National Dengue incidence. Solid lines represent the “direct” forecast of the national incidence and dashed lines are the “aggregate” approach: the national forecast being the aggregate of the states forecasts. The legend in the top left panel denotes the eight different SARIMA models. The left, middle and right panels display the: mean absolute error (MAE), mean relative absolute error (MRAE), and ratio of the MAE of the models to the MAE of the NULL model.

For Thailand (results not shown), the direct method performed slightly better than the aggregate method for one-, two-, and four-month windows. In contrast, for Brazil, the aggregate model consistently outperformed the direct model across all prediction horizons. Moreover, the differences between the two was significant. For example, the MAE for the best model, over a two-month horizon decreased from 0.51 to 0.39, or 23%.

### Effects of a Reporting Time Delay

We found that building reporting delays into the data available to make the forecasts resulted in a significant degradation in forecasting accuracy, with predicted peaks drifting later in time, becoming lower in magnitude, and with an overall reduction in the total number of cases (Fig. 8). Although this could be inferred from the ever-increasing size of the forecast window (e.g., Fig. 3), a clearer picture results from explicitly building the delay into the statistical model. For this purpose, we chose the 1-month-3-year autoregressive model ((1, 0, 0)(3, 0, 0)^12^) and forecast one-month forward incidence profiles assuming: no delay, a one-month delay, a two-month delay, and a three month delay (Fig. 8). When data are provided timely, one month predictions are remarkably accurate, with both the amplitude and phase of the peak being well predicted. The most serious discrepancy lies in the one month phase lag that the model produces, being unable to anticipate the increase or decrease in the gradient of the curve. Moving progressively from one- to three-month delays, the predicted amplitude of the peak decreases and the peak drifts later into the season. For a three-month delay, the model forecasts a peak delayed by *∼* 5 months, and completely misses the second smaller peak (at least for the case of Thailand during the 2015-2016 season). Similar problems arise in other locales (SI, Fig. SI S6-S8).

**Fig 8.**
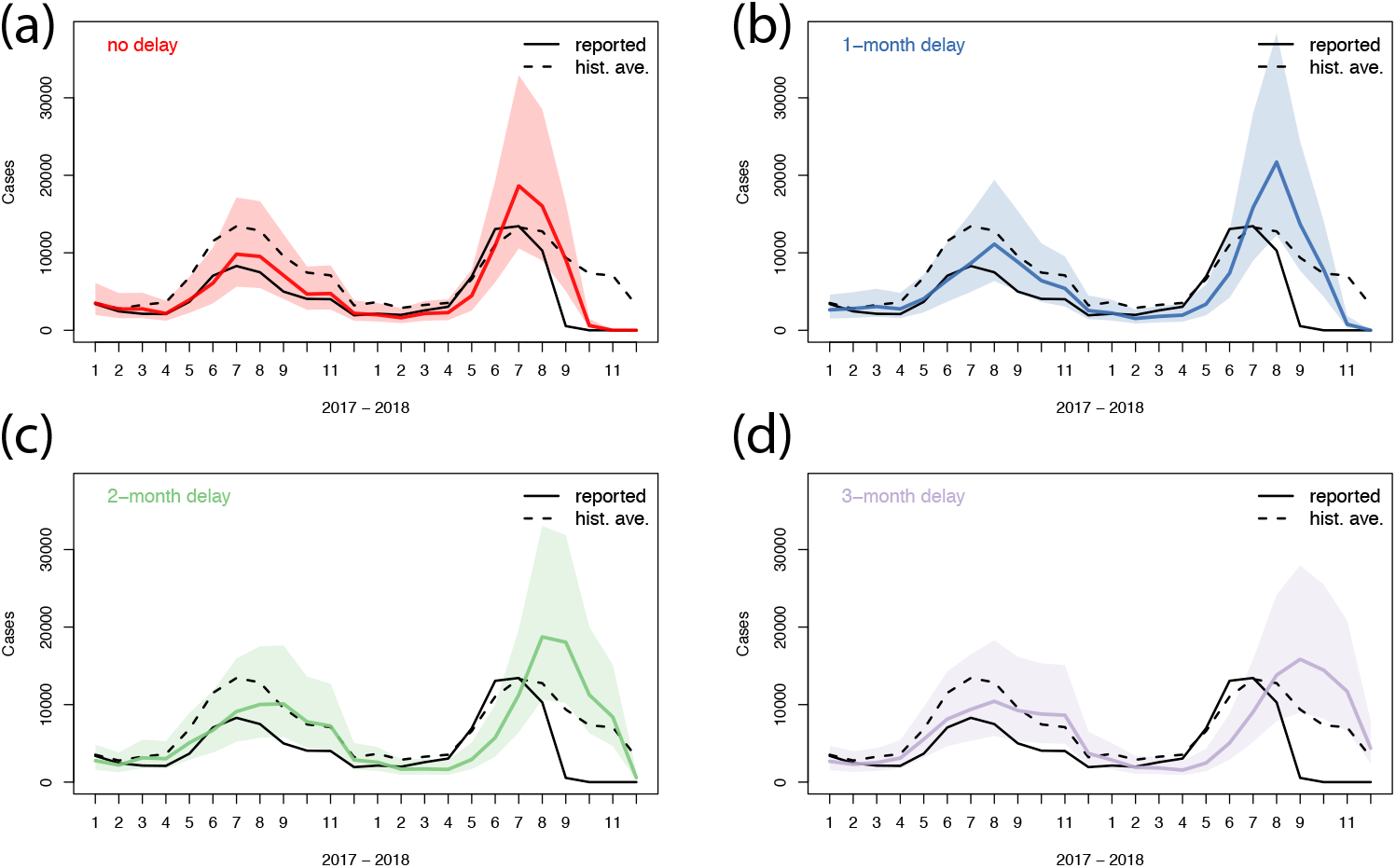
Effect of time delay for Thailand national Dengue incidence: (a) no delay; (b) one-month delay; (c) two-month delay; and (d) three-month delay. The curves show: Reported cases (black solid), historic monthly average (dashed), forecast (red, blue, green, and purple, respectively), and 5/95% confidence bounds (lighter shading).

Singapore incidence, being reported weekly, allows us to explore the degradation in forecasting accuracy at a higher temporal resolution: Assuming sequentially: 0, 1, 2, …, 9-week delays in the reporting of cases, we can estimate the decrease in accuracy of forecasts over a two-month period (Fig. 9). As was found for monthly data, across virtually all datasets, the effect of increasing reporting delay are as follows. First, the peak drifts later in time. That is, peaks are predicted to occur later. Second, the amplitude of the peaks are eroded. Third, the total number of cases are predicted to be less than is observed. Interestingly, the differences between the observed and predicted pattern at eight weeks from weekly data is qualitatively the same as the differences at two months using monthly data. Thus, the forecast is not obviously sensitive to the resolution of the data used, only the duration of the interval being forecast. In summary then, a balance between the need to have timely data and the desired window over which one would like to be able to make accurate forecasts.

**Fig 9.**
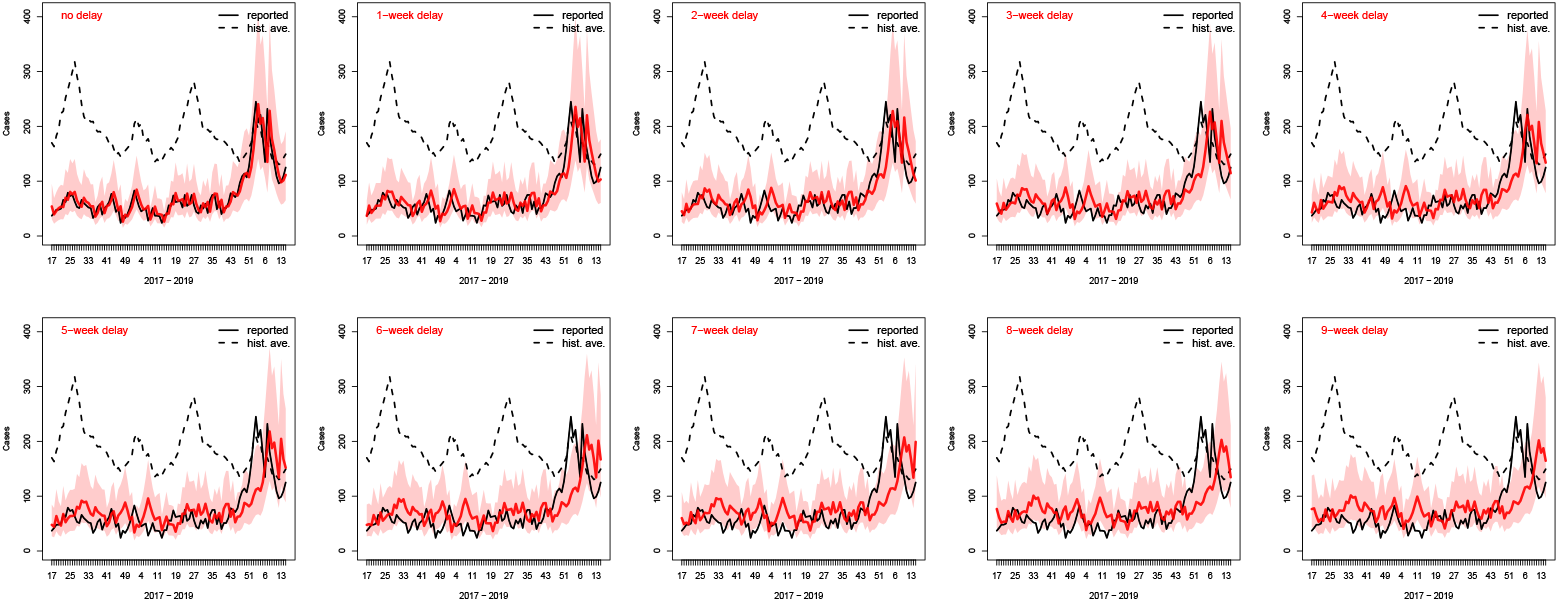
Effect of time delay for Singapore weekly Dengue incidence with delays of zero to nine weeks. The black curve shows the data, the red curve is the model forecast, and the shaded area marks the 5/95% confidence intervals. The dashed black curve is the historic weekly average value.

## Discussion

In this study, we have used the publicly available data to developed a general time series framework for making near-term forecasts of Dengue incidence in six different countries, at multiple spatial resolutions, with and without climatic data. We found that SARIMA models generally outperform a ten-year historic average “null” model out to the longest time horizon considered (4 months). Additionally, we showed that modest improvements are generally seen when including precipitation covariate data in the model. We demonstrated the importance of timely reporting in terms of the degradation in the quality of the forecast. Our code can be downloaded from a GitHub repository and, with little effort, adapted to other Dengue data sources or even different pathogens entirely. Additionally, we are continually updating the sources from which we collect and publish these data.

We explicitly defined a null historical model in this study to act as our baseline or reference model against which to test the SARIMA models. This model can be thought of as a reflection of implicit non-numerical forecasts that are made by public health professionals. We suggest that this null model allows us to assess which time-series models could be considered to consistently outperform intuitive experience-driven opinion.

Previous studies have applied statistical, and, in particular, SARIMA techniques to study and predict the near-term evolution of Dengue incidence in different locations. These include Brazil [18, 21],and São Paulo in particular [15], Mexico [6], Singapore [22], and Thailand [23]. These (and other related) studies, while broadly consistent, did not fully agree with the results presented here. For example, one investigation concluded that climate data did not improve the predictive power of the SARIMA model for Dengue incidence in Mexico [6], while another, using data from Thailand, found that it was a crucial component [23]. It may be that the precise SARIMA techniques that were implemented led to these conflicting conclusions, or that there are intrinsic differences in the shape of incidence curves for the disease in these two vastly separated regions.

Our results and conclusions are generally consistent with those of Johannson et al. [6], who found that both the (1, 0, 0)(2, 1, 0)^12^ and (1, 0, 0)(3, 1, 0)^12^ models performed well across most Mexican states. The interpretation was that both short-term positive auto-correlation and also long-term negative seasonal auto-correlation are driving factors in forecasting Dengue. Our analysis of the Mexico data (extending further in time than the previous study) suggests that both (1, 0, 0)(3, 0, 0)^12^ and (1, 0, 0)(3, 1, 0)^12^ models perform well. The long-term negative seasonal auto-correlation effect could be understood intuitively from a visual inspection of the multi-year time series: Following an anomalously large season (usually produced by the introduction of a new serotype) the next seasons will have ever smaller peaks, until a new large peak returns. Thus, in spite of the fact that the SARIMA model does not include serotype information, that knowledge is communicated to the forecast through the incorporation of seasonal information.

Our inclusion of climatic co-variate data generally improved forecasts more so than has been reported elsewhere for individual populations [6]. Specifically, its inclusion either made the forecasts more accurate, or at least did not degrade accuracy. Intuitively, this makes sense: Dengue transmission is driven by mosquito density, which in turn is strongly dependent on the presence of standing water. That previous results were more ambiguous probably reflects limitations from studying only a single location. Our analysis across multiple continents promotes a more statistically-compelling conclusion. Nevertheless, even here, the gains are not substantial (e.g., the addition of climatic co-variates does not improve the accuracy such that a three-month forecast with climatic data is as accurate as a two-month forecast without climatic data). It is likely that the month-scale resolution of the incidence, which constrains the resolution of the climatic data, is leading to such a weak effect. We anticipate that daily or even weekly data would provide greater accuracy gains, although we note that this was not the case for the Singapore data analyzed here. We believe that the lack of any improvements in forecast accuracy when we included precipitation as a covariate for Singapore stems from the vastly different epidemic curves from one year to the next.

The impact of reporting delays on potential situation awareness during a dengue season is substantial. With seasons typically lasting *∼* 2 *−* 5 months, even a delay of one month can render forecasts of little immediate benefit. However, when that delay becomes two months or more, even four month forecasts have dramatically reduced accuracy, to the point that historic averages now outperform even the best SARIMA model. Currently, Dengue latency ranges from several weeks (e.g., Singapore and Sri Lanka) to potentially many months (e.g., Mexico data are typically published once per year). Our results provide a strong argument for prioritizing the availability of up-to-date incidence reports, particularly during the early phases of an outbreak. However, an important caveat to reducing the latency in data availability is a possible trade-off with accuracy. The retrospective analysis performed here did not account for any reporting errors; however, data published in near-real-time would be subject to a range of errors, including under-reporting, mis-diagnosis of symptoms, and/or false test results. Later, these would presumably be adjusted as the sources of error are corrected (e.g., [24]).

In addition to providing a comprehensive analysis of these different Dengue data sources, a second goal of this study was to create and make available a data repository of the data-streams, updated in near-real-time, together with the R code necessary to reproduce the plots discussed here (see https://github.com/predsci/Dengue or http://www.predsci.com/dengue). Additionally, forecasts for all available countries, states, provinces, and districts for which we have data are available online [25].

While time series models are valuable from a forecasting point of view, they are limited scientifically in that they do not provide any mechanistic insight into the underlying biological processes. Thus, an obvious next step is to use a similar assessment framework to test forecasts made with dynamic models, which, ultimately should provide better forecasting capabilities, whilst at the same time, providing clues about the relevant processes. We anticipate that while statistical models may outperform mechanistic approaches on short horizons (*<* 4 months, say), mechanistic models will likely provide better predictions at longer time horizons [24].

## Materials and Methods

### Data

Data used in this study were obtained from a number of sources. For each dataset, we developed a data-scraping algorithm that would periodically visit the appropriate website, identifying any new reports of incidence and appending them to our current database. Specifically: (1) Brazil data were retrieved from both the national SINAN system [26] and the department of health in Rio de Janeiro [27]; (2) Mexico data were obtained from the Mexico Directorate of Epidemiology [28]; (3) Singapore data were obtained from the Singapore Ministry of Health [29]; (4) Sri Lanka data were obtained from the Epidemiological Unit of the Sri Lanka Ministry of Health [30]; and (5) Thailand data were retrieved from the Thai Ministry of Public Health’s weekly epidemiological surveillance reports [31].

Precipitation data were obtained from NOAA using the NCEP-DOE Reanalysis II dataset. This dataset is provided on a 2.5^*?*^-resolution global grid [32]. For each region, we constructed population-weighted averages of precipitation using an overlay of Global Administrative Maps (GADM) [33] and SocioEconomic Data and Applications Center (SEDAC) [34] population density maps.

## Models

We used a suite of SARIMA models implemented in R and provided as part of the “stats” package. In general terms, the models can all be described as:

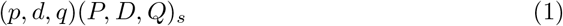

where, for the non-seasonal components (*p, d, q*), *p* specifies the number of lags, *d* represents the degree of differencing, and *q* is the order of the moving-average model. The next three parameters (*P, D, Q*)^*s*^ specify the “seasonal” aspects of the model, with *s* being the length of the season (12 months or 52 weeks in our study) and *P, D*, and *Q* equivalent to the non-seasonal parameters. As an example, consider the model (1, 0, 0)(1, 0, 0)^12^. This corresponds to a model that includes a non-seasonal autoregressive (AR) term of order one, a seasonal AR(1) term, no differencing, no moving average terms and a seasonal period of 12 months: this is an AR model with predictors at lags of 1, 12, and 13 months.

Following Johansson et al. [6], we limited *p* to values of 1, 2 or 3, *P* to values 0, 1, 2, or 3, and *d* and *D* to either 0 or 1. Thus, lags were limited to 3 months (*p*) or three years (*P*). Finally, both *q* and *Q* were fixed at 0. Thus, in general, there were up to 32 models under consideration. It is worth making a few comments on the intuitive interpretation of this modeling approach. Model selections with values of *p* = 3 and *P* = 3, for example, suggest that past values from three months and three years ago are important for forecasting purposes. The parameter *d* informs the model how much differencing should be applied; a process that may be important if the stationarity assumption is not well met. A previous study of Mexico Dengue fever, which looked at the effects of climate covariates in the SARIMA analysis, and, in particular, temperature showed that while SARIMA models benefited from the additional information, Seasonal SARIMA models did not [6], their contribution is, at best, marginal. Interestingly they found that at the state level, the inclusion of climatic covariates actually worsened the forecasts.

As a covariate, we used precipitation, and, in particular, added it as a lag term of 0, 1, or 2 to the most promising SARIMA models (*p* = 1, *P* = 1, 2, 3, *d* = *q* = *D* = *Q* = 0, and *s* = 12). Under real-time forecasting conditions, we would need a forecast of this information for three months ahead. Although this data were available for most of the duration of this retrospective study, the most likely source of such data for a prospective forecast would be monthly historic averages. Thus, these were computed and used here.

## Data Availability

Anyone who wishes to share, ruse, remix, or adapt this material must obtain permission from the corresponding author.

## Acknowledgments

The authors thank Nathan Sherburne for his assistance in scraping the data analyzed in this study. The work reported here was performed under the auspices of the U.S. Department of Defense’s Defense Threat Reduction Agency.

## Supporting information

**Fig S1.**
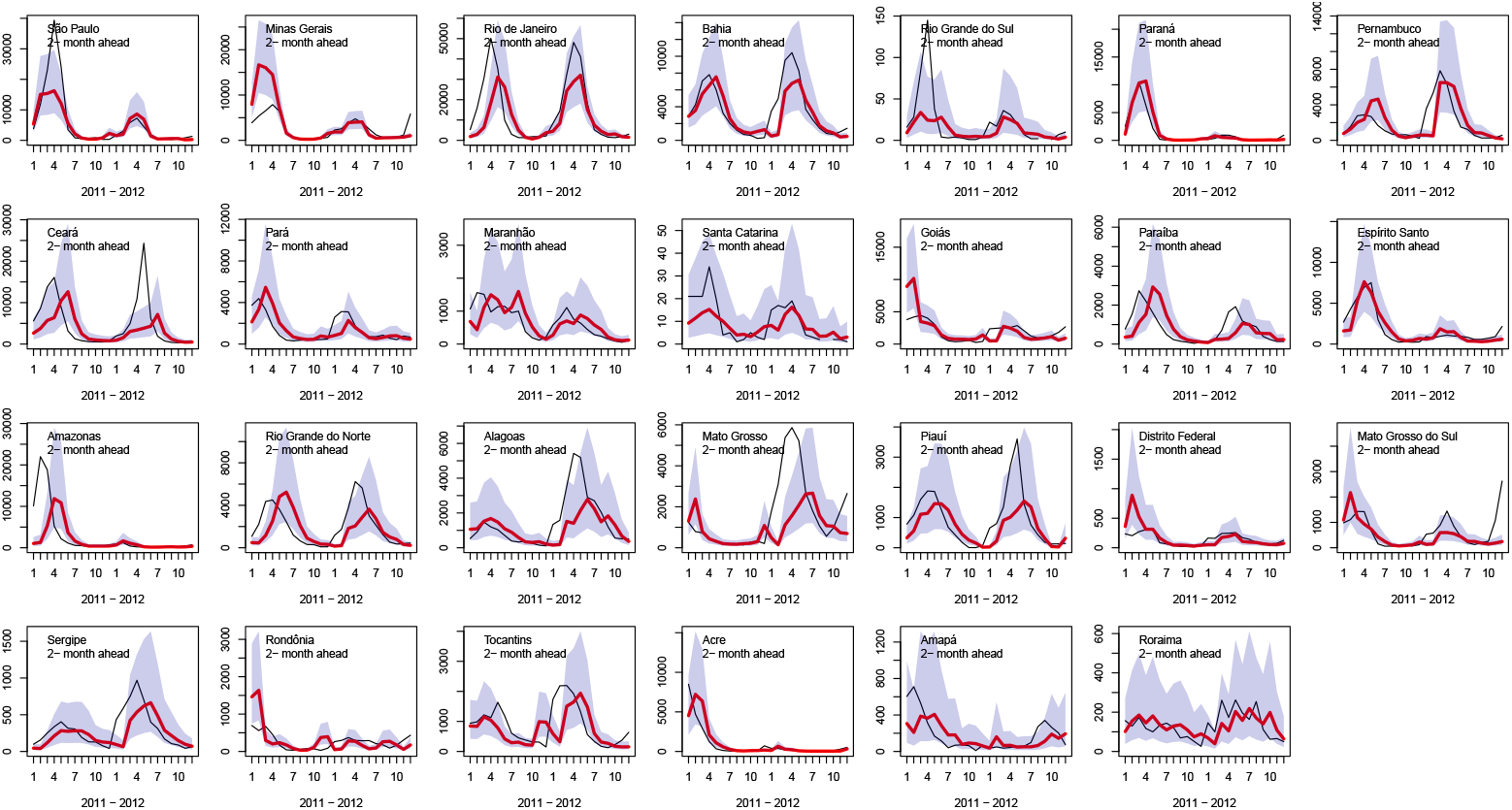
As Fig. 4 but for two-month predictions.

**Fig S2.**
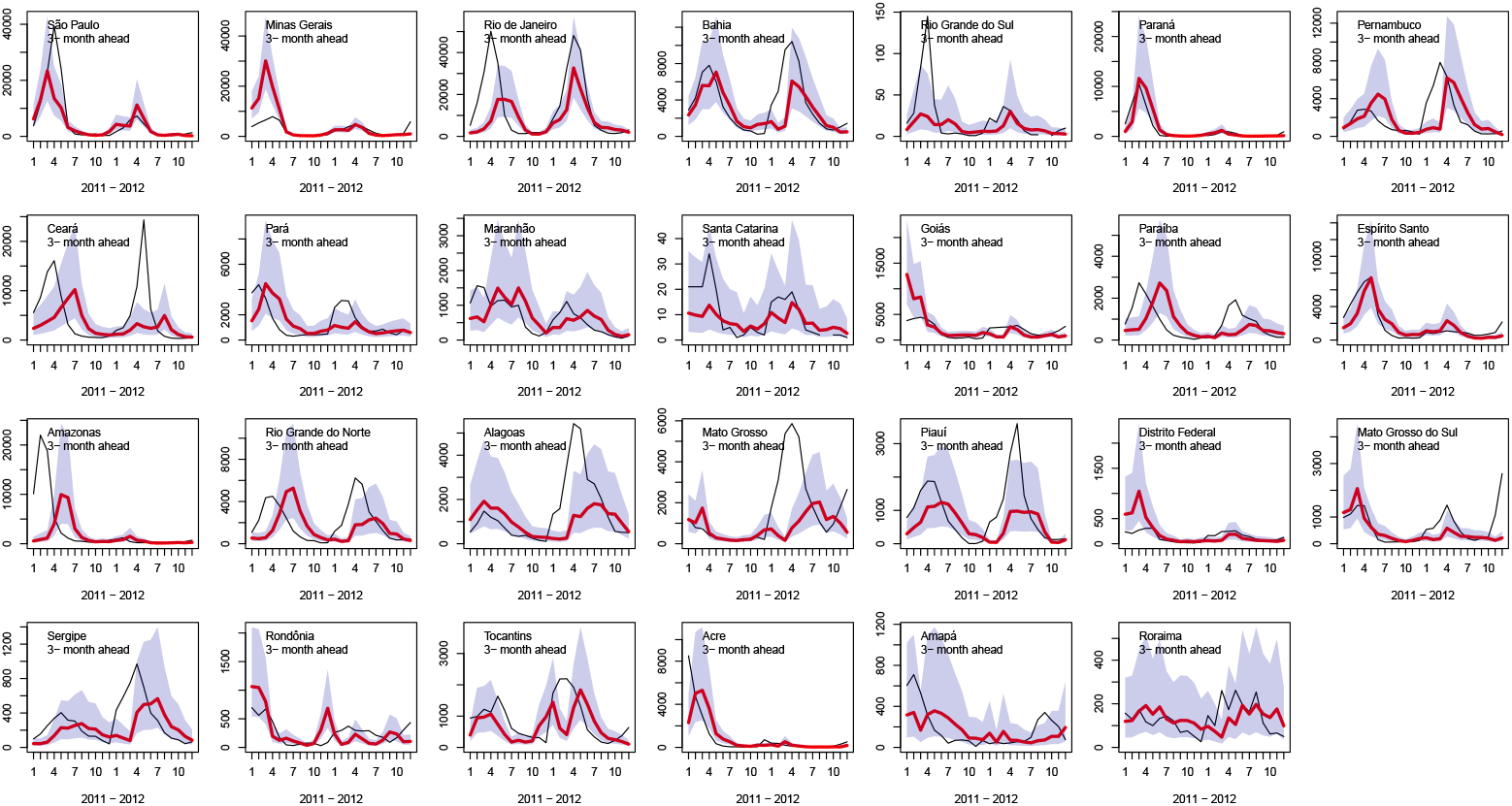
As Fig. 4 but for three-month predictions.

**Fig S3.**
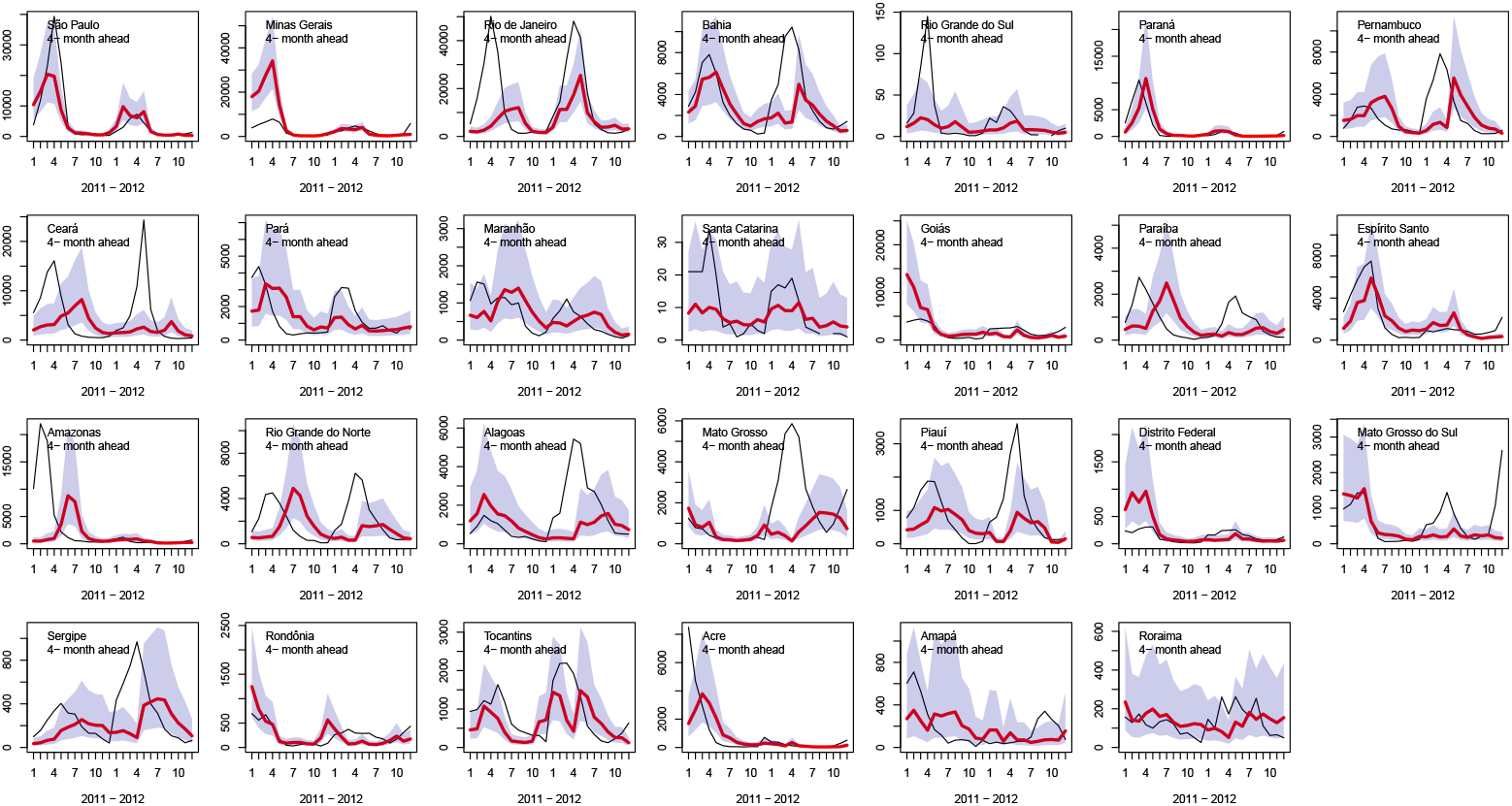
As Fig. 4 but for four-month predictions.

**Fig S4.**
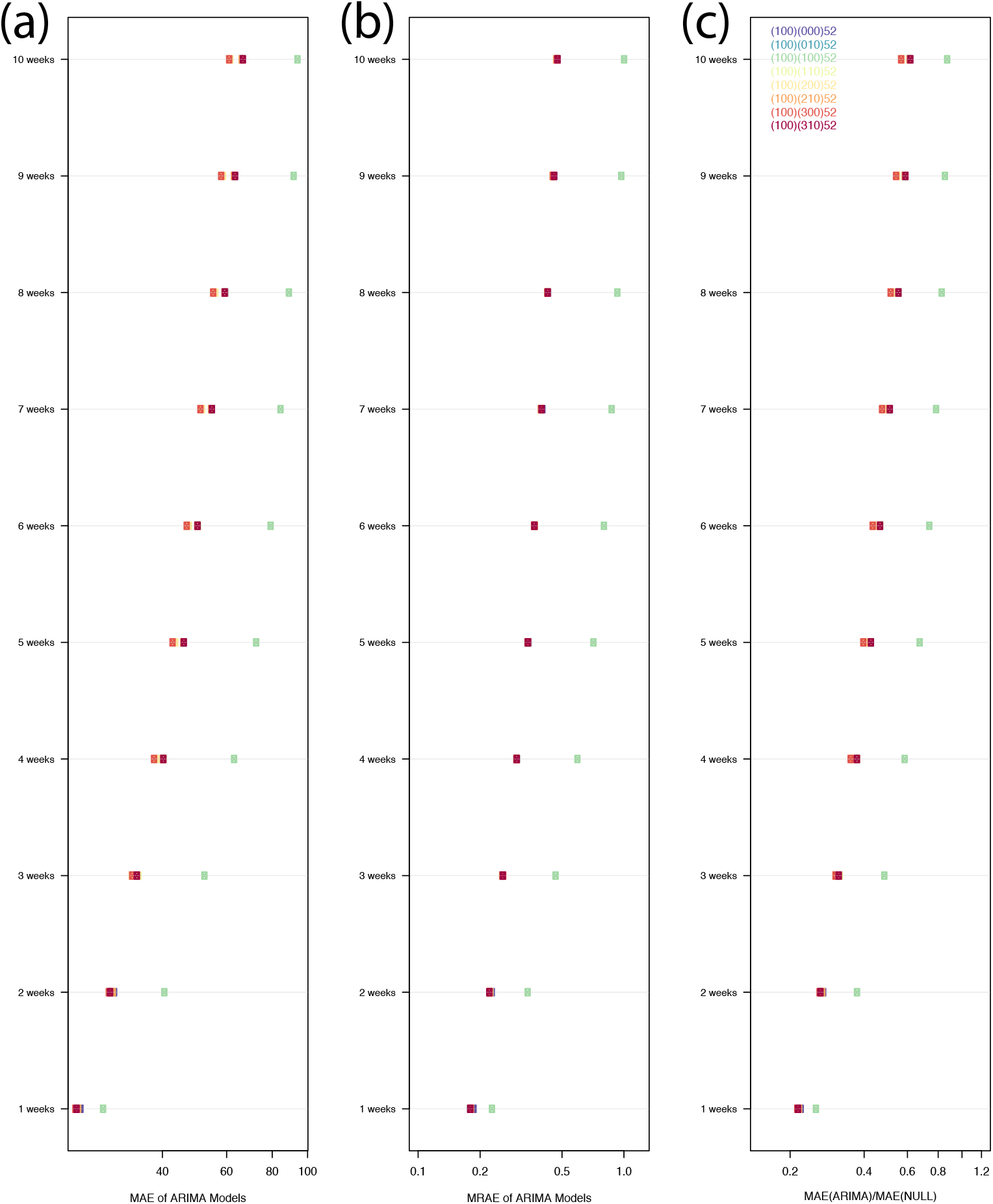
As Fig. 2, but for Singapore weekly data with forecast horizons ranging from 1 to 10 weeks.

**Fig S5.**
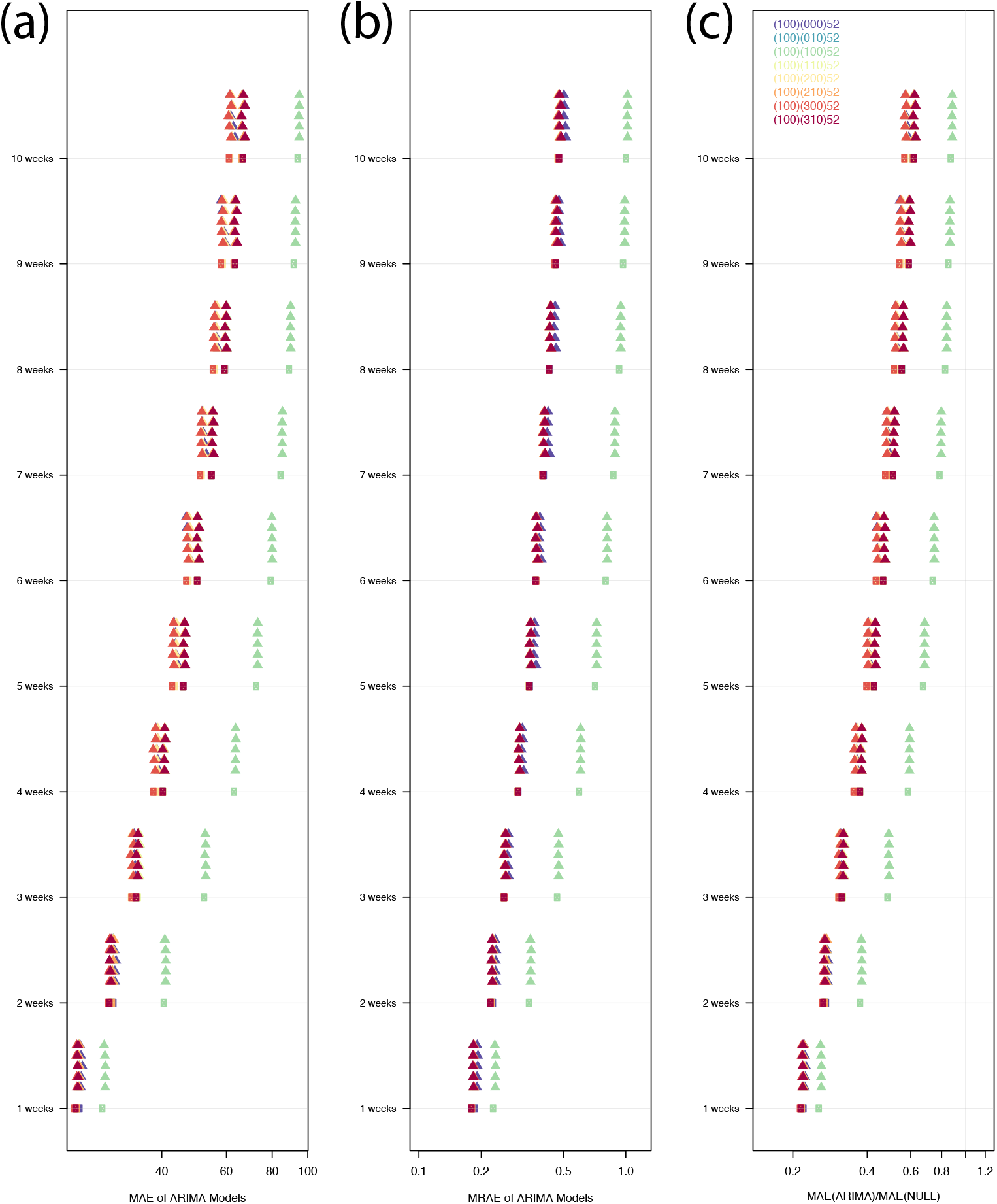
As Fig. S4 but including precipitation as a covariate.

**Fig S6.**
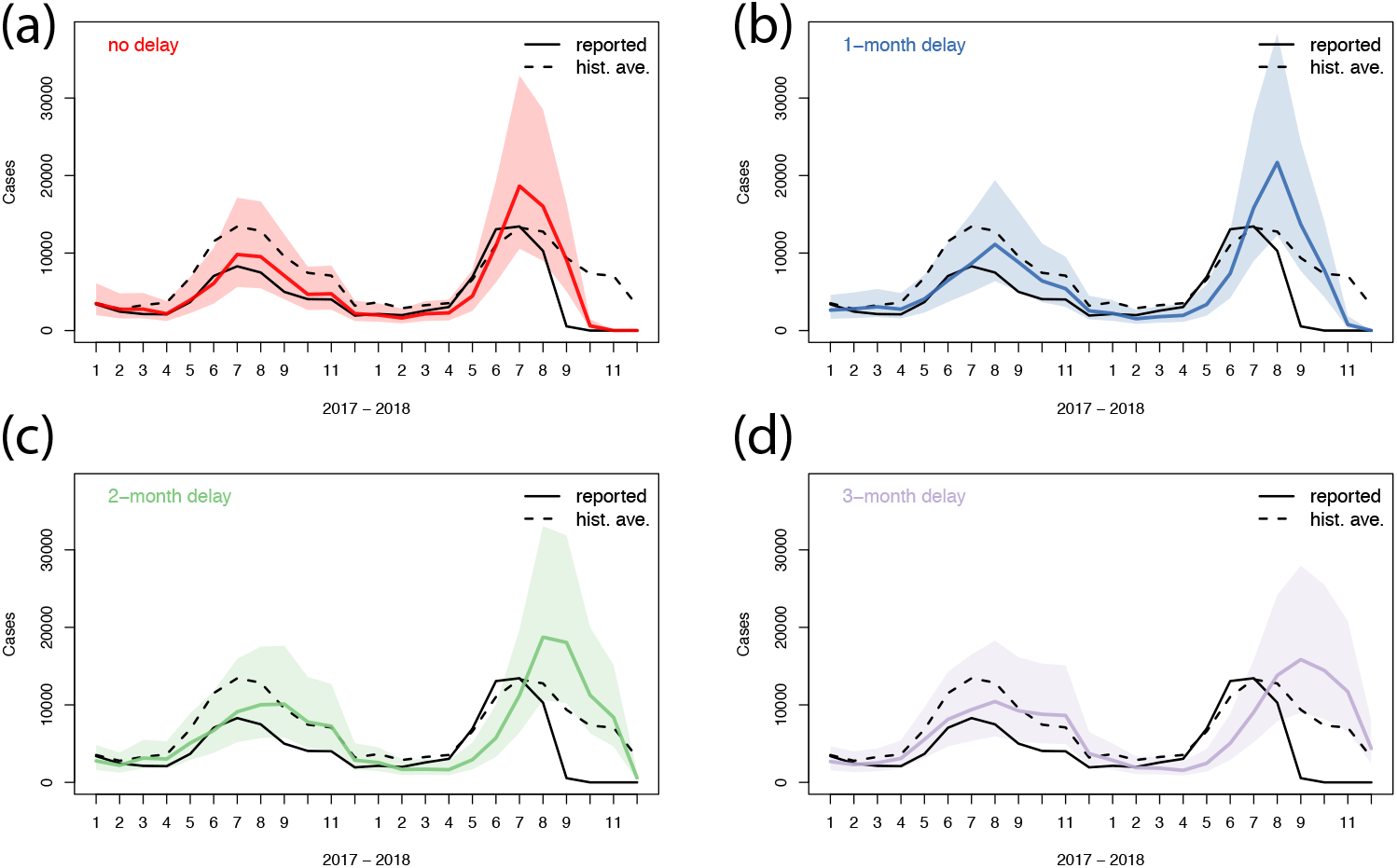
Effect of time delay for Thailand national Dengue incidence: (a) no delay; (b) one-month delay; (c) two-month delay; and (d) 3-month delay. The curves show: Reported cases (black solid), historic average (dashed), forecast (red, blue, green, and purple, respectively), and confidence bounds (lighter shading)

**Fig S7.**
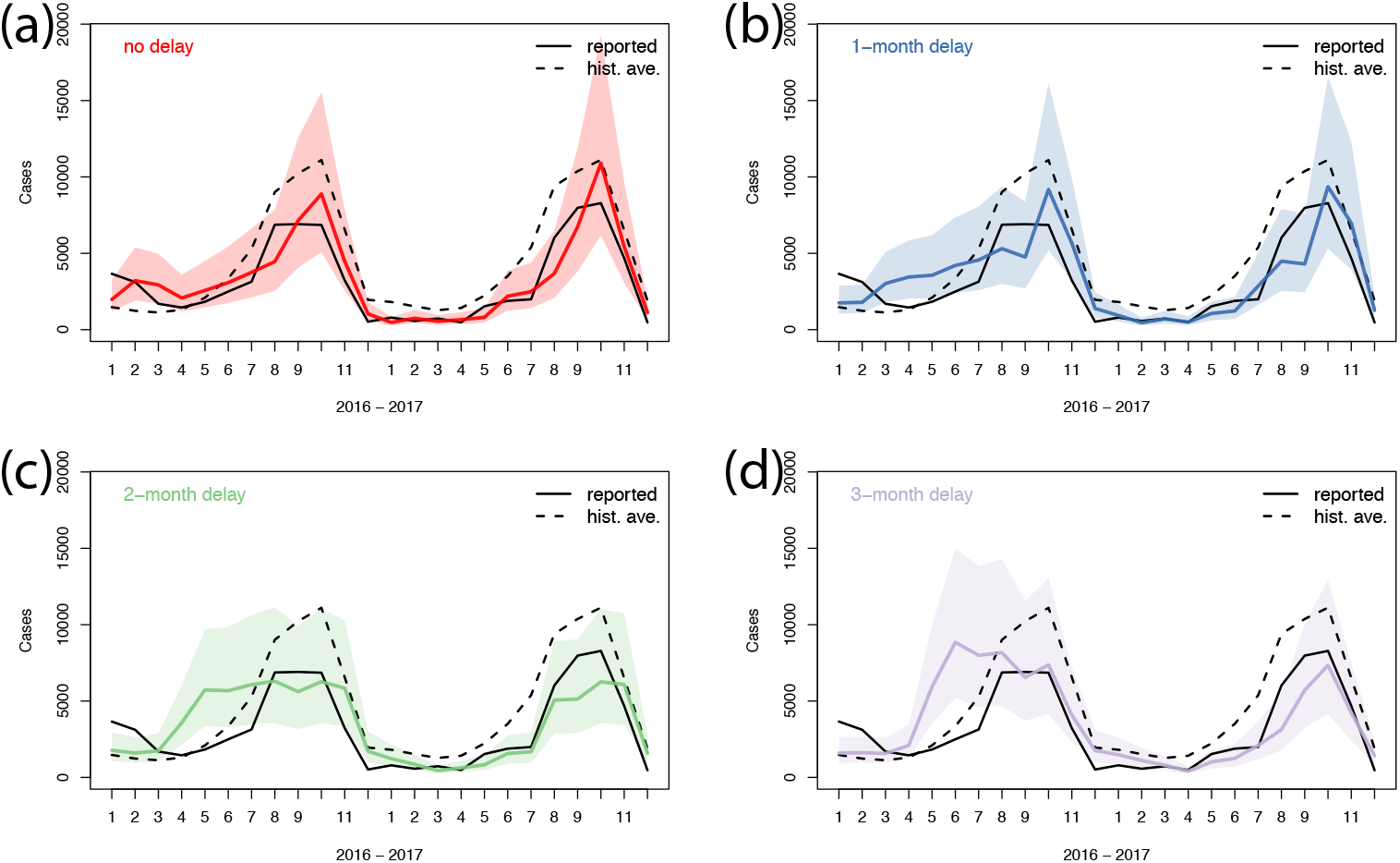
As Fig. S6 but for Mexico national incidence.

**Fig S8.**
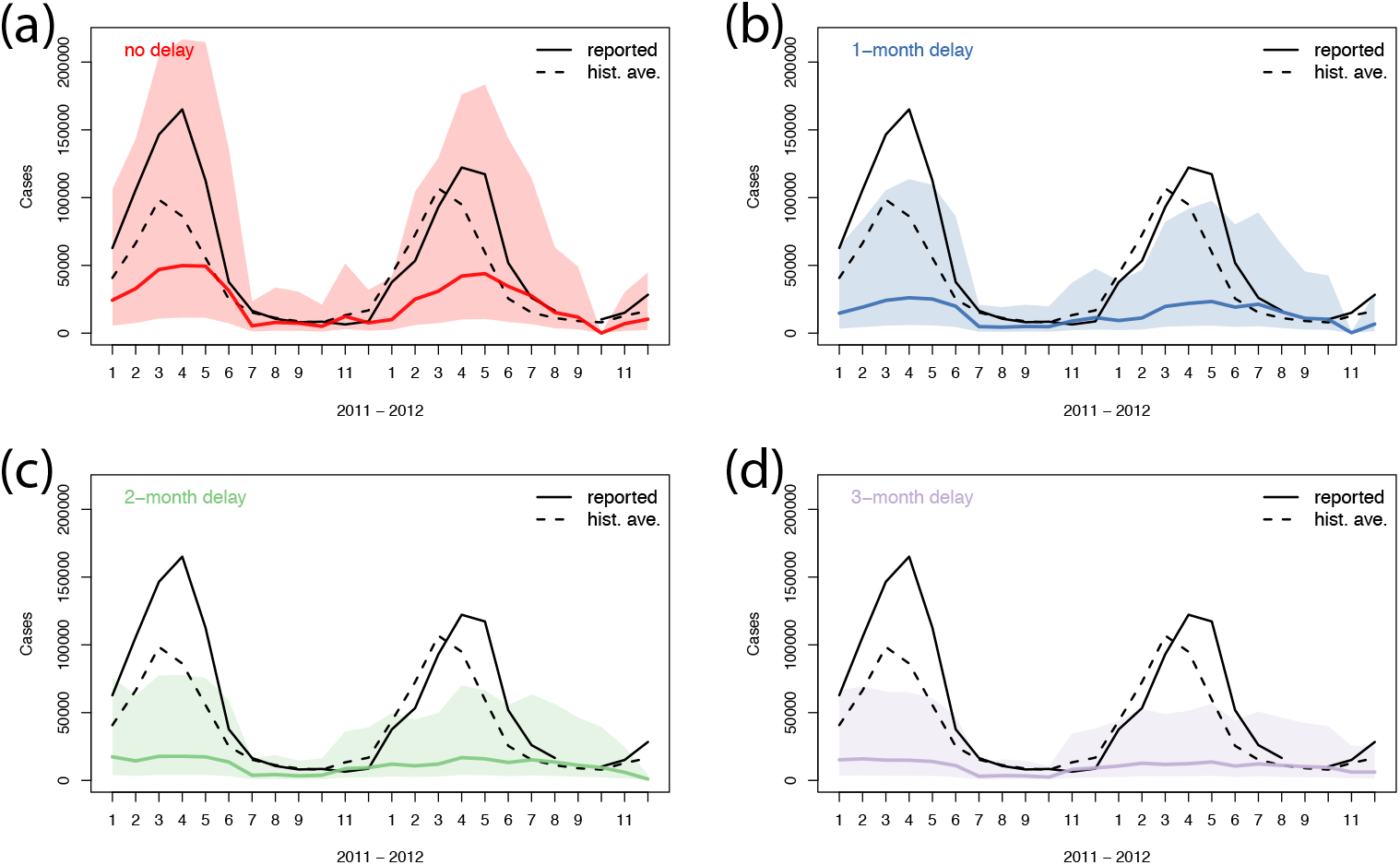
As Fig. S6 but for Brazil national incidence.

